# “It was brutal. It still is”: A qualitative analysis of the challenges of bereavement during the COVID-19 pandemic reported in two national surveys

**DOI:** 10.1101/2021.12.06.21267354

**Authors:** Anna Torrens-Burton, Silvia Goss, Eileen Sutton, Kali Barawi, Mirella Longo, Kathy Seddon, Emma Carduff, Damian JJ Farnell, Annmarie Nelson, Anthony Byrne, Rhiannon Phillips, Lucy E. Selman, Emily Harrop

## Abstract

The COVID-19 pandemic has been a devastating, mass bereavement event characterised by sudden unexpected deaths and high levels of disruption to end-of-life, grieving and coping processes, as well as social life more broadly. We analysed qualitative free-text data from two independent UK-wide online surveys to describe in depth the experiences of 881 people bereaved during the Covid-19 pandemic using. We analysed the data in two phases, conducting an inductive thematic analysis and then applying Stroebe and Schut’s Dual Process Model (DPM) (1999; 2010) as an analytic lens to further contextualise and interpret the data. The DPM identifies loss-oriented and restoration-oriented coping processes between which grieving people naturally oscillate. Loss-oriented coping involves coming to terms with the death and lost relationship, while restoration-oriented coping involves adapting to new ways of life. We identified six main themes: troubled deaths (guilt, anger and unanswered questions); mourning, memorialisation and death administration; mass bereavement, the media and the ongoing threat of the pandemic; grieving and coping (alone and with others); work and employment; and support from the health and social care system. Examples of loss-oriented stressors included being unable to visit or say goodbye, the sudden and traumatic nature of many deaths, and restricted funeral and memorialisation practices. Associated reactions were feelings of guilt and anger, and problems accepting the death and starting to grieve. Examples of restoration-oriented stressors and reactions were stressful death-related administration and severely curtailed social networks, support systems and social/recreational activities, which impacted people’s ability to cope. Study results demonstrate the exceptionally difficult sets of experiences associated with pandemic bereavement, and the utility of the DPM for conceptualizing these additional challenges and their impacts on grieving. Our analysis builds and expands on previous use of the DPM (Stroebe and Schut, 2021) in explicating the impact of the pandemic on bereavement. We make recommendations for statutory, private and third sector organisations for improving the experiences of people bereaved during and following this and future pandemics.

## BACKGROUND

The COVID-19 pandemic represents a devastating, mass bereavement event, accompanied by profound levels of social and economic disruption on a global scale. The unexpectedness of most COVID-19 deaths, lack of access to and physical contact with relatives at the time of death and restrictions surrounding funerals are highly distressing for bereaved relatives, with potential long-term impacts on the grieving process (Hanna et al. 2021; Pearce et al 2021). Lack of access to usual support networks and severe societal disruption compound these risks for everyone bereaved during the pandemic (Goveas et al 2020; Harrop et al 2021; Stroebe and Schut 2001, 2021). For these reasons, and based on parallels with other mass bereavement events, researchers have predicted increases in the proportions of people experiencing prolonged grief disorder (PGD) and other mental health problems (Eisma et al. 2020, Menzies et al 2020).

A review by Stroebe and Schut (2021) systematically examined and categorised these types of pandemic-related bereavement circumstances into loss-oriented stressors and reactions and restoration-oriented stressors and reactions, as conceptualised in their Dual Process Model (1999;2010). The model describes normal grieving as bereaved people oscillating between focussing on the loss of the deceased person (loss-orientated coping, e.g., grieving) and negotiating the practical and psychosocial changes to their lives that occur as a result of the bereavement (restoration-orientated coping, e.g., forming new roles/identities/relationships). Stroebe and Schut suggest that this natural oscillation process is likely to be disrupted in the context of the pandemic, requiring people to modify their loss-orientated and restoration-orientated activities and behaviours, with potential longer-term impact on bereavement outcomes. Examples of pandemic-specific loss-orientated stressors include lack of emotional and practical preparation time, traumatic deaths (including being unable to say goodbye and suboptimal care), profoundly altered funeral practices and lack of social/cultural recognition of the loss. Stroebe and Schut suggest reactions to these stressors may include guilt, shame, anger and loneliness. Examples of restoration-oriented stressors are loss of work, disrupted living arrangements and family dynamics, erosion of coping resources, disruption to routines and loss of pre-crisis ways of life. Restoration-oriented reactions include anxiety and diminished sense of control or purpose, as well as feelings of vulnerability and insecurity. However, whilst their review usefully identifies and conceptualises these different types of factors in relation to the model, they acknowledge that most of the included articles were either expert opinion pieces or reviews of pre-pandemic studies published early on in the pandemic, with very little COVID-19 specific empirical data considered (Stroebe and Shut 2021).

Results from recently published pandemic studies provide evidence for some of these bereavement circumstances and their impacts. One preliminary study (Eisma et al. 2021) confirmed higher levels of PGD for people bereaved by COVID-19 and unnatural deaths (i.e. accidents), compared to natural bereavement (i.e. deaths from chronic illness). Another study observed higher levels of functional impairment for all deaths during COVID-19 compared to pre-pandemic times, but no differences between COVID-19 and other types of deaths (Breen et al. 2021). Quantitative and qualitative studies have identified difficult experiences of end-of-life care, such as lack of communication and contact with healthcare staff and patients prior to the death (Becque et al 2021, Hanna et al 2021, Mayland et al 2021, Neimayer and Lee 2021; Selman et al 2021). Where bereaved people struggled to make sense of such experiences, ‘disrupted meaning’ was found to cause functional impairment and dysfunctional grief symptoms (Breen et al. 2021). The distress caused to family members by visiting restrictions, missed opportunities to spend time with and say goodbye to their dying family member, and feelings of frustration at poor communication from healthcare staff is documented in the qualitative results of one of these studies. Such experiences were often described as traumatic, and accompanied by feelings of sadness, guilt, anxiety and feeling ‘cheated’ (Hanna et al 2021; Mayland et al 2021). High levels of loneliness, social isolation and emotional support needs have also been observed amongst bereaved participants in the current study, in conjunction with difficulties accessing both informal and formal sources of bereavement support (Harrop et al 2021; Selman et al 2021c).

Overall, however, there remains a lack of rich qualitative evidence on the many different aspects of pandemic bereavement experiences, the meaning and consequences of these experiences for bereaved family members, and which considers existing bereavement theory as a lens for understanding these experiences. Based on qualitative analysis of free text-data from two UK-wide surveys, we describe in detail the lived experiences of people bereaved during the COVID-19 pandemic, aiming to generate a rich understanding of the challenges that they have faced. Following initial inductive analysis, we assessed the explanatory ‘fit’ between our data and the DPM and conducted further directed analysis using the DPM as an analytic lens (Stroebe and Shut 1999). We consider our findings in light of the DPM and make recommendations for end-of-life care and bereavement support.

## METHODOLOGY

### Study designs and aims

The qualitative data that we analysed was collected in two national surveys:

#### BeCOVID

The Bereavement during COVID-19 (BeCOVID) Study aimed to investigate the grief experiences, support needs and use of bereavement support by people bereaved during the pandemic by any cause of death. The study includes a longitudinal survey with three timepoints: baseline (28th August 2020 to 5th January 2021) and two follow-up surveys approximately 7 and 13 months after the death of the loved one. We report on the qualitative free-text data from the baseline survey.

#### COPE

The UK COVID-19 Public Experiences (COPE) Study was a longitudinal mixed-methods study which aims to understand the public’s experiences and responses to the COVID-19 pandemic and government policy during the first 12 months of the UK outbreak (Hallingberg et al 2021; Phillips et al 2021). Three online surveys were conducted over a 12-month period: baseline in March/April 2020, 3-month follow-up in June/July 2020, and 12-month follow-up in March/April 2021. We report on qualitative data on bereavement experiences collected as part of an optional module in the 3-month follow-up survey (20th June to 20th July 2020).

### Participants

#### BeCOVID

Participants were aged 18 years + with the ability to consent; family or close friend bereaved since social-distancing requirements were introduced in the UK (16/03/2020); death occurred in the UK (n = 711).

#### COPE

Participants were adults aged 18+ years living in the UK; data were included from individuals willing to answer the optional module from the 3-month follow-up survey, who had experienced a bereavement between 1/03/2020 and 20/07/202 when the survey closed (n = 499/7,043).

### Survey development

#### BeCOVID

An open web survey (see Harrop et al. 2021) was designed by the research team, which includes a public representative (KS), with input from the study advisory group. It was piloted, refined with 16 public representatives with experience of bereavement and tested by the advisory group and colleagues. Open and closed questions covered end-of-life and grief experiences, and perceived needs for, access to and experiences of formal and informal bereavement support.

#### COPE

The optional bereavement module was developed by authors (EH, RP) to enable the cohort of participants who had experienced a bereavement to tell us about their experiences. Participants were asked if they experienced any of the following challenges: limited contact with loved one at end-of-life, unable to say goodbye properly, restricted funeral arrangements and social isolation following the bereavement. An open-text box was provided, and participants invited ‘to write about these or any other experiences. For example, you may like to tell us about how your bereavement affected you and how you think support could have been improved’.

### Study procedure

#### BeCOVID

The baseline survey was administered via JISC (https://www.onlinesurveys.ac.uk/) and was open from 28th August 2020 to 5th January 2021. It was disseminated via social and mainstream media, voluntary sector associations and bereavement support organisations, including those working with ethnic minority communities. Organisations helped disseminate the voluntary (non-incentivised) survey by sharing on social media, webpages, newsletters, on-line forums and via direct invitations to potential participants. For ease of access, a web link to the survey was posted onto a bespoke study-specific website with a memorable URL (covidbereavement.com). Two participants completed the survey in paper format.

#### COPE

The voluntary (non-incentivised) baseline COPE survey through which participants enrolled to the study was disseminated using a multi-faceted sampling method based on convenience sampling, snowballing, and purposive sampling via social media (Facebook®, Twitter® and Instagram®). The study was also advertised via HealthWise Wales (HWW), a national population survey and research register of residents who live or receive healthcare in Wales. The 3-month follow-up survey that included the bereavement questions was administrated using Qualtrics.com and disseminated between 20^th^ June and 20^th^ July 2020 to participants who completed the baseline survey (Hallingberg et al 2021, Phillips et al 2021). Towards the end of this survey, participants were given the option to answer an optional module on their experiences of bereavement during the pandemic (since 1st March 2020). Respondents’ demographic data and corresponding answers to the bereavement module were extracted from the main data set (n = 7,048) to enable focused analysis of bereavement experiences.

The initial sections of both surveys requested informed consent and details data protection.

### Data analysis – both data sets

Free-text survey responses were analysed using inductive thematic analysis, involving line-by-line coding in NVivo V12 and identification of descriptive and analytical themes (Braun and Clarke 2006). A preliminary coding framework was developed based on a sample of survey responses from both surveys (ATB, EH, ES). The framework was revised and applied in an iterative process (EH, ES, SG, KB, ATB), moving between the data and the analytical concepts to develop codes and themes grounded in the data. This involved independent double coding of 10% of both datasets, regular discussion and cross-checking within the study team and review of final themes by the wider qualitative team.

Following initial inductive analysis, we assessed the explanatory ‘fit’ between our themes and the DPM and conducted further deductive analysis using the DPM as analytic lens (Stroebe and Shut 1999). We decided to apply this model retrospectively due to its empirical credibility and wide application in pre-pandemic bereavement studies (Stroebe and Shut 2010), its explanatory ‘fit’ with many of our themes, and the value that it brings for conceptualising and advancing our understandings of the many challenges of pandemic bereavement. We mapped our themes to the lists of pandemic-specific Loss and Restoration Oriented Stressors and Reactions developed by Stroebe and Shut (2021), identifying new examples in our data (Tables 3 and 4, Supplementary file 1).

## RESULTS

### Sample Characteristics

#### BeCOVID

711 participants who had been bereaved completed the survey (Table 1), with 626 (88%) providing free-text comments. 88.6% of participants were female (n=628); the mean age of the bereaved person was 49.5 years old (SD = 12.9; range 18-90). The most common relationship of the deceased to the bereaved was parent (n=395,55.6%), followed by partner/spouse (n=152,21.4%). 56% of deaths (n=399) were non-covid related. 10 % of people (n = 72) had experienced more than one bereavement since 16th March 2020.

**Table 1.**
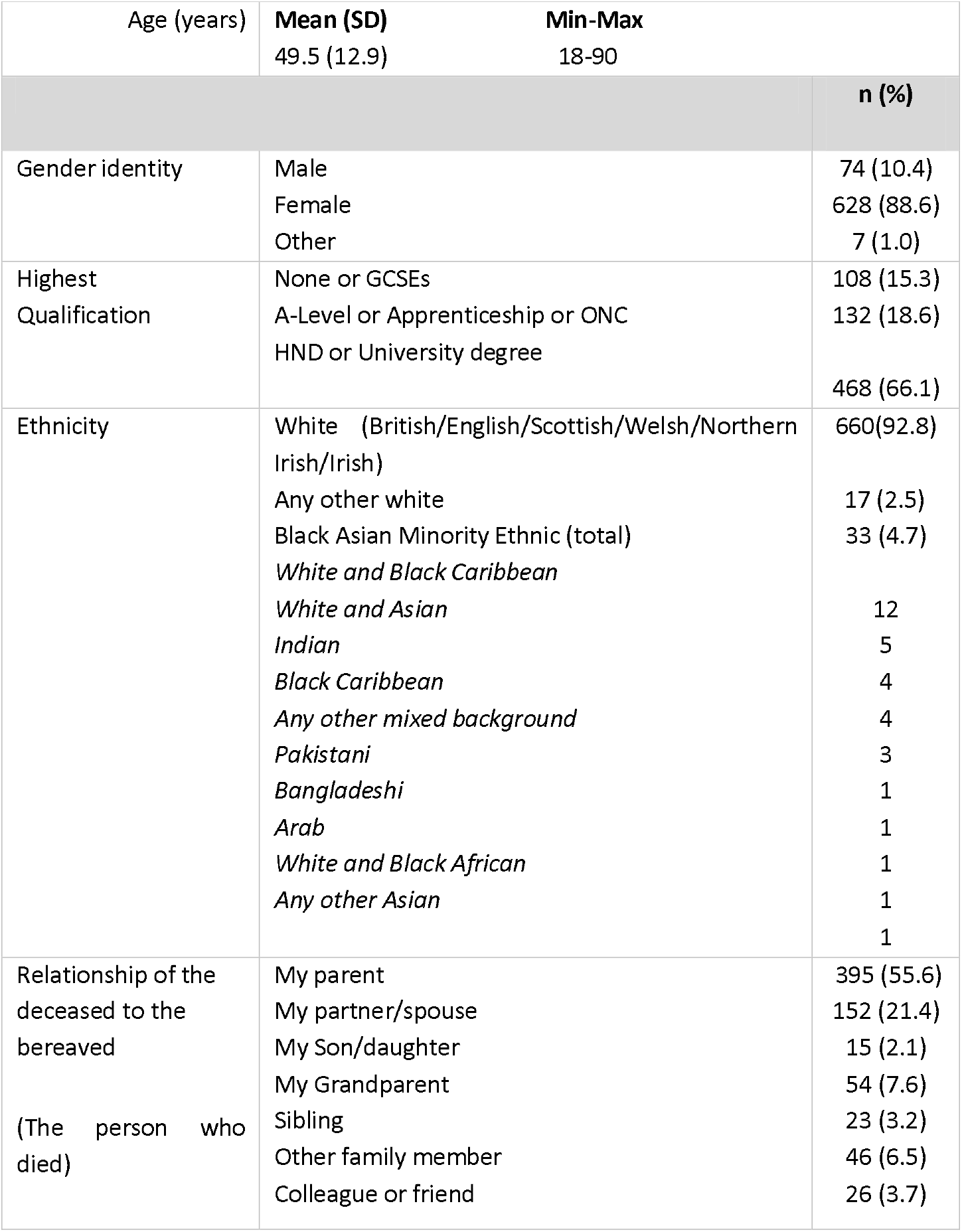
Participant demographics from BeCovid data n = 711)

#### COPE

499 bereaved participants completed the bereavement survey module (Table 2), with 51% (n = 255) providing free-text comments. 75% of participants were female (n = 376); the most frequent age range of the bereaved person was 61 to 70 years (n= 146, 29.3%). The most common relationship of the deceased to the bereaved was other family member (e.g., grandparent, aunt/uncle) (n=254, 50.9%), followed by a friend (n=115, 23%). 59% of deaths were reported to be non-COVID related (n=295). 8% of participants (n=40) reported to have experienced more than one bereavement since 1st March 2020.

**Table 2.** Participant demographics from COPE data (n = 499)

|  |  | **n (%)** |
| --- | --- | --- |
| Age | 18 to 30 | 32 (6.4) |
|  | 31 to 40 | 55 (11.0) |
|  | 41 to 50 | 90 (18.0) |
|  | 51 to 60 | 100 (20.0) |
|  | 61 to 70 | 146 (29.3) |
|  | 71 to 80 | 64 (12.8) |
|  | 81 + | 12 (2.4) |
| Gender | Male | 121 (24.2) |
|  | Female | 376 (75.4) |
|  | Prefer not to say |  |
| Highest Qualification | Usual high school qualifications at age 16 (e.g. GCSE, O-level) | 56 (11.2) |
|  | Usual high school qualifications at age 18 (e.g. AS level, A level) | 41 (8.2) |
|  | A college or university diploma or degree | 230 (46.1) |
|  | A higher degree or professional qualification (e.g. a Doctorate or Masters level) | 121 (24.2) |
|  | None of these qualifications | 21 (4.2) |
|  | Other | 24 (4.8) |
|  | Rather not say | 4 (0.8) |
| Ethnicity | White(English/Welsh/Scottish/Northern Irish/British) | 482 (96.6) |
|  | White- other |  |
|  | Mixed/multiple ethnic groups | 9 (1.8) |
|  | Other | 3 (0.6) |
|  | Rather not say | 1 (0.2) |
|  |  | 2 (0.4) |
| Relationship of deceased to the bereaved<br>(the person who died) | My partner | 9 (1.8) |
|  | My parent | 19 (3.8) |
|  | My sibling | 27 (5.4) |
|  | My son/daughter | 59 (11.8) |
|  | Other family member (e.g. grandparents, aunts/uncles) | 254 (50.8) |
|  | Friend | 115 (23.0) |
|  | Colleague | 13 (2.6) |
|  | Neighbour | 14 (2.8) |
|  | Other | 28 (5.6) |

### Themes

The combined data-sets include free-text responses from a total of 881 participants. We identified six major themes across the two studies, presented with sub-themes in Figure 1. The major themes related to: troubled deaths (guilt, anger and unanswered questions); mourning, memorialisation and death administration; mass bereavement, the media and the ongoing threat of the pandemic; grieving and coping (alone and with others); work and employment; and support from the health and social care system. In the quotes below, ‘BRID’ indicates the participant reporting number from the BeCOVID study and ‘CRID’ from COPE. These IDs are only known to members of the research team.

**Figure 1:**
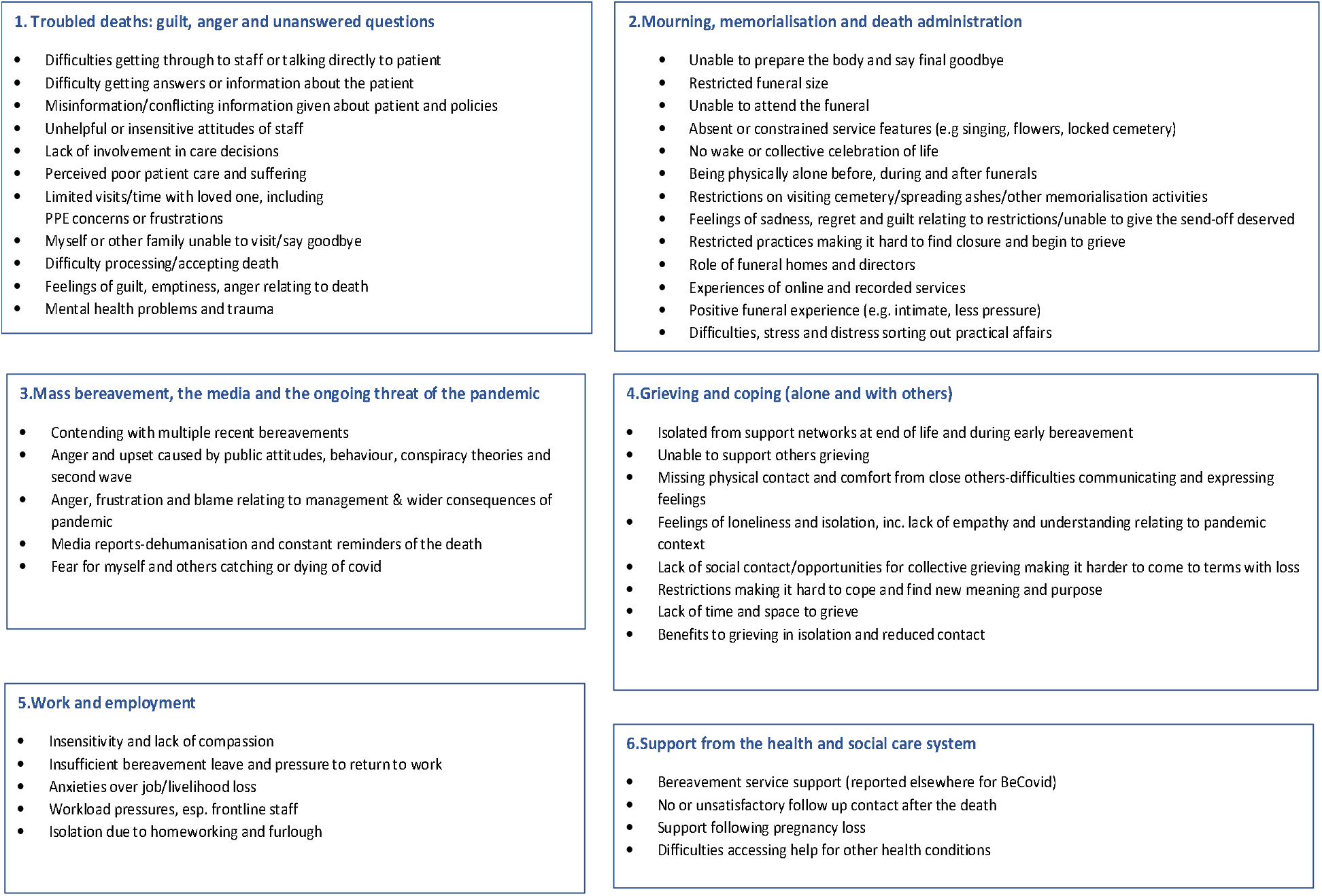
Main themes and sub-themes

#### Troubled deaths: guilt, anger and unanswered questions

The end of life and death of a relative or friend is often a difficult time; however, the pandemic has added significant complications to these experiences, exacerbating feelings of distress and grief. People commonly described communication problems with healthcare providers, including difficulty contacting staff by telephone and getting information about their relative/friend, misinformation concerning the patient’s condition and hospital policies, perceived staff insensitivity, and a lack of involvement in care or treatment decisions. Some people also raised concerns around the quality of care provided to their relative/friend.

> *‘My wife was sent home from hospital to die, in a situation where I feel she could have received medical care. She was left without adequate care for three days over a weekend as they discharged her too quickly for a care package to be arranged. When the package was implemented, she was already critically ill but no one actually told me. No one told me how poorly she was, and this left me in a situation where she died before I said goodbye. I am struggling to deal with this.’ (Bereaved husband, CRID704)*

Infection control measures denied many people the opportunity to spend time with their dying relative/friend, especially in hospital and care-home settings. This was especially the case in the earliest months of the pandemic when visits were prohibited, but also occurred later on when misinformation or untimely communications meant that they missed the death, despite officially being allowed to visit when the patient was deemed “end of life.” Participants described mixed experiences of communicating with their sick relatives through video calls on phones or iPads. Whilst some valued the contact that this gave them, others described frustrated attempts caused by poor internet connection, problems with equipment and being unable to hear the conversations properly; this exacerbated people’s sense of heartbreak and frustration that they could not physically visit. Amongst those who were able to visit, many described difficult experiences and anxieties related to wearing personal protective equipment (PPE) and unclear guidance on the use of this. For some, the need to wear masks and gloves caused significant upset as it interfered with their desire for final physical contact. People bereaved early on in the pandemic also described their upset at the lack of protection available around the hospital site (e.g. hand sanitizer) and their related worries over contracting and spreading the virus either to the sick patient or other family members.

> *‘I feel resentment towards the pandemic for robbing me of the last 5 weeks of my mum’s life. Whilst I was able to visit her the morning she passed away, holding her hand through a nitrile glove & being unable to kiss her one last time due to the full PPE head mask was difficult to cope with, as much as I cherish being allowed to see her in her final few hours.’(Bereaved daughter, BRID523)*

This lack of contact at the time of death, often combined with the sudden and unexpected nature of COVID-19 deaths, intensified the sense of loss and pain. Death experiences were described as ‘traumatic’, depicted as ‘nightmares’ and were associated with severe anxiety and feelings of panic. The inability to visit or say goodbye left many people with feelings of intense sadness and guilt that they could not be there to comfort and support their loved one. Participants reported a sense that they ‘failed them’ and worried that they may have felt abandoned. This was particularly true for participants whose relatives died in hospitals and care homes, with relatives of those who lived in care homes describing extended periods of separation. Where relatives had dementia further upsets were described relating to problems using virtual methods of communication to stay in contact, and the potential distress caused to them by widespread PPE use and limited physical contact. For some, the lack of contact prior to their death meant that the loss felt less ‘real’ and harder to accept. For those acting as the main point of contact with care settings, the pressures of being the ‘messenger’ and having to provide answers to other family members and friends added to their stress and emotional burden, particularly when tensions developed.

> *‘Mum was in hospital for a period of time then back to the nursing home where she died. I hadn’t seen her at all since March due to being in a home but was allowed to see her once before she died and after she had died. It was upsetting not to be able to speak to her and communicate with her properly before she died. Feeling of guilt that I wasn’t with her.’ (Bereaved daughter, CRID637)*

Negative emotions such as anger at being unable to visit, at the care that their loved ones received or treatment decisions that were made, further affected relatives’ ability to process and reconcile their feelings surrounding the death. Having unanswered questions or doubts over how or why they died, and feelings that the deaths may have been avoided by the earlier introduction of infection control measures or, for non-covid deaths, continued access to treatment, compounded this anger and upset.

> *‘My partner had her treatment (as part of a clinical trial) stopped because of COVID. They said the risks if she got COVID were too high. However, she died anyway. She had one trip to hospital where the doctor failed to send her out of hospital with essential antibiotics. Because carers weren’t allowed into hospitals I didn’t know this had happened and she didn’t know what should have happened*… *I am not complaining, I believe everyone was trying their best and sometimes it wasn’t enough… however because it’s no one’s fault there is nowhere to place the anger, the complex emotions above and beyond ‘normal’ bereavement are too huge to process. This needs to be recognised.’ (Bereaved partner, BRID487)*

#### Mourning, memorialisation and death administration

Disruption to the bereavement process continued following the death. People were unable to visit their loved ones in the chapel of rest or funeral homes to say a final goodbye, and people bereaved by COVID-19 described the distress caused by ‘closed coffin’ requirements. Families were unable to complete rituals, such as dressing and preparing their loved one before burial. These were seen as ‘final acts of love’ and their impossibility was reported to have directly impacted their grieving.

> *‘We couldn’t dress her, do her hair or put little things in the coffin or wake her properly. She was always well turned out and she always took great care of her hair. We couldn’t do any of those final acts of love for her.’ (Bereaved daughter BRID296)*

Restricted funeral practices caused further upset to grieving family members, especially when attendance was limited to ten or fewer people and taken-for-granted practices such as readings and singing were prohibited. Families had to make difficult decisions regarding who attended, while travel restrictions meant that people from outside the local area were unable to attend or be closely involved with funeral arrangements. Close family members and friends who were shielding or self-isolating were also prevented from attending. Many people described their upset at not having their wider family and friends at the funeral, being unable to sing chosen songs or conduct religiously appropriate ceremonies. They expressed general sadness that their loved one could not have the “send-off” they deserved. People also commented on the added distress caused by having to travel to and from funerals alone, sit apart from friends and family and being unable to comfort or be comforted by them.

> *‘It was an horrendous experience not being able to hug and cry with family members, not being able to give loved one a proper send off, the whole thing was a harrowing experience.’ (Bereaved relative, CRID145)*

The supportive role and efforts of funeral directors were positively noted by many participants. Descriptions were given of innovations and adaptations made to memorialisation practices and services. Virtual streaming of funeral services received mixed appraisals; some people found this ‘stark,’ whereas others were deeply appreciative that this technology was available. Examples were also given of local communities lining the streets to pay their last respects. A minority of participants appreciated the intimacy and reduced stress associated with smaller, quieter funerals. However, for most, being unable to host conventional services or wakes, share stories and celebrate the life of their loved ones was deeply upsetting, and made it difficult to find closure and begin to grieve. Consequently, many people felt that their loved one had not been remembered as they would have liked, and that their grief and bereavement was unreal and ‘on hold’.

> *‘Funeral was small and still feel we haven’t properly said goodbye, so many of her friends often ask when we will be able to do a memorial service feels as though her life has gone and not been fully recognised for the person she was.’ (Bereaved sister, BRID137)*

This disruption and upset to memorialisation practices continued beyond the funeral. Restrictions to cemetery visits and being unable to scatter ashes in chosen resting places caused further distress. The need for dedicated memorial spaces and remembrance activities was also mentioned, particularly in the face of COVID-scepticism in the media and among a minority of the general public. The hopes of some people bereaved early in the pandemic to have ‘proper celebrations’ after lockdown restrictions eased were fading as the pandemic progressed. There was a growing realisation that this may never happen due to ongoing restrictions and a sense that the moment had passed, adding to their sadness and regret.

> *‘I wish there was a dedicated memorial in our town, maybe in a park or near the town hall where we can sit and remember. I’ve found it incredibly hard to listen to the rubbish being spouted on tv by various ‘celebrities’ saying it’s all a lie and a hoax. My dad’s death is not being used to boost their flagging career. I’m sick of listening the narrative coming from the government.’ (Bereaved daughter, BRID98)*

People also described how the practical and administrative aspects of dealing with their bereavement had been made more difficult by the disruption caused by the pandemic to professional services. These included obtaining death certificates, arranging funerals over the phone, informing financial and other agencies of the death, and selling and vacating the houses of those who died. Long administrative delays and difficulties getting in touch with the right people/agencies was highly stressful for family members already grieving. Unresolved life insurance claims were a further source of anxiety and the emotional toll of dealing with the property and possessions of the deceased was noted.

> *‘Since his death have had so many problems trying to get things sorted out and it has been a nightmare of missed calls and writing letters to wait months for a reply. This has been a sad soul destroying journey which I wouldn’t wish upon anyone else, it hurts and is still hurting and confusing questions still remain unanswered.’ (Bereaved wife, BRID167)*

#### Mass bereavement, the media and the ongoing threat of the pandemic

The distress and grief complications caused by restrictions on visiting and mourning practices were further compounded by the ongoing nature and wider societal consequences of the pandemic. Around one in ten participants had suffered multiple bereavements since the start of the pandemic, reflecting on how overwhelming an experience it had been to lose more than one close contact in such a short space of time, whilst others recalled the added strain that they felt as a result of recent pre-pandemic losses. One participant described the additional distress caused by the disproportionate effects of the pandemic on minority ethnic communities, and the societal inequalities that it has highlighted.

> *‘It has been hard as a Black person, seeing how many Black, Asian and minority ethnic citizens have been impacted by the Coronavirus, along with White communities of course. The pandemic has laid bare how many health and care inequalities exist in British society. It is my hope that in future, and as a result of the unavoidable public enquiry, health and care outcomes for disadvantaged groups improve dramatically in coming decades.’ (Bereaved son, BRID731)*

Participants described the dehumanising effect of being bereaved at a time of mass bereavement and the negative impact of daily death tolls announced in the media. Some people reflected on how this widespread trauma left them feeling unable to openly grieve or seek support. Feelings of anger and alienation were caused by perceptions of government incompetence in handling the crisis, conspiracy theories questioning the pandemic shared in social and mainstream media, along with members of the public and officials disregarding social-distancing requirements and regulations. Such behaviours were seen not only as disrespectful and exacerbating the threat posed by the disease, but also as undermining opportunities for their families to grieve under more ‘normal’ conditions.

> *‘The enormity of the loss of life confronting the UK has left me with the feeling that l cannot openly grieve the loss of my friend or formally seek support. One friend suggested l should take comfort from the fact so many others are also grieving, l found this comment distressing. I felt silenced and shut down.’ (Bereaved friend, BRID153)*

Constant coverage of the pandemic also meant that participants’ grief and trauma felt inescapable, with daily reminders of the circumstances surrounding the death. Participants bereaved by COVID-19 described anxiety relating to the continued threat of the virus and the looming prospects of a ‘second wave’.

> *‘I’m struggling with COVID being around still, thus one of the only things that is talked about every single day. I try to avoid the news to refrain from being constantly reminded of my loss. I’ve had to put a barrier up so high to not let insensitive comments directed unintentionally at me, affect me. I’ve had to remind myself that this is affecting the whole world on different levels and to try not to take anything personally.’ (Bereaved daughter, BRID215)*

Fear of catching or spreading COVID also had a significant impact on people’s adjustment and ability to cope following the bereavement, particularly amongst those bereaved by COVID-19. People found it difficult to prioritise their own grief when worrying about themselves or other family members contracting the virus, particularly if they were clinically vulnerable themselves. This fear surrounding the disease also made it difficult for people to go about their daily lives. Many participants described anxieties about going shopping, socialising, returning to frontline jobs, or their children returning to school.

> *‘I fear that the same will happen to me as I’m [clinically vulnerable] and have two [children] and I don’t want them to go through losing their mum in the same way. I’m terrified of them going to school and getting infected. How can I grieve when I’m terrified and trying to protect them.’ (Bereaved daughter, BRID15)*

#### Grieving and coping (alone and with others)

Many people were separated from their usual support networks during lockdown conditions. Grieving processes were impacted by being unable to meet up to remember their relatives/friends and to support each other in their grief. Social isolation made it more difficult to grieve and to come to terms with their loss. Feeling distant from and lacking physical contact with close others made their grieving feel ‘artificial’; it was more difficult to share memories, discuss feelings openly and begin to come to terms with and process their shared grief and loss. It was also more difficult to access or provide the emotional support that was needed, and many participants reported loneliness. People described great upset and frustration at being unable to provide support to other family members or to the family of friends who had passed away. They also had concerns over not troubling others at a time of universal suffering and hardship and felt that (non-bereaved) others could not understand what they were going through due to the exceptional nature of pandemic bereavement.

> *‘As a group of friends we haven’t been able to help each other and hold each other through this difficult time. It came as a massive shock, and I feel so lost. I want nothing more than to be around my friends.’ (Bereaved friend, CRID756)*

A minority of people perceived benefits to grieving in private, without having to manage difficult social situations. Some benefited from close contact with their immediate family, whereas others found comfort and support through virtual methods of communication. However, for others, their constant closeness to their immediate family during lockdowns made it harder to process their feelings. Some felt that they had less space and time to grieve properly due to family responsibilities, which were intensified by home-schooling and in some cases the mental health needs and difficulties of other family members. Feelings of isolation and emotional distress were also acute when family members had pre-existing strained relationships or complex dynamics or where new tensions and conflicts emerged surrounding the death/post-death period.

> *‘I found being on lockdown with my wife and children made it extremely hard to get in touch with the grieving process - 24 hours a day, 7 days a week of responsibilities to others did not give me the space I needed to even think about my loss properly, and the strange atmosphere of lockdown made it hard to pinpoint what was going on for me emotionally around the loss of my dad.’ (Bereaved son, BRID144)*

Many participants grappled with a loss of meaning and purpose. This was not only in relation to their lost relationship and associated roles, activities and life-plans, but also with respect to difficulties finding new purpose and hope in the context of pandemic restrictions and suppression of taken-for-granted ways of life.

> *‘At first you have a purpose. The funeral. Sorting things out. Afterwards and I think due to the pandemic I have little purpose or meaning in life. I focus for work but I don’t see the point. The smallest thing rocks me.’ (Bereaved partner BRID391)*

People reported feeling overwhelmed and unable to make important life decisions in response to changing circumstances caused by the pandemic. Participants with pre-existing mental health problems described how restrictions made it much harder for them to manage their condition. People struggled with not being able to leave the house to visit friends or to engage in activities or hobbies. They were unable to volunteer or attend church or to experience respite from their situation. Many participants described a need to keep busy and to try to maintain as much of a ‘normal’ life as possible, although this was very difficult during periods of lockdown.

> *‘As a remote worker, once I went back to work it’s been harder to deal with the isolation and maintain a work/life balance. I’ve been a carer for a parent with cancer since I turned 18 [….] and I’m not used to having time for myself and to indulge in hobbies, and with the current restrictions it’s almost impossible to take up a hobby so I can find myself working late to fill the time.’ (Bereaved daughter, BRID697)*

#### Work and employment

A number of problems relating to workplaces and employment were reported, in particular perceived insensitivity and a lack of understanding and compassion amongst managers and colleagues. People described feeling pressured to return to work before they felt ready, and managers expressing disappointment when doctors notes were used to extend their bereavement leave. Some felt that they were judged negatively for poorer performance, made to feel that they were not coping, and that their employers’ expectations for their recovery were unrealistic. Others thought that by taking time off they would be ‘giving up’. A number of participants reported needing time off further into their bereavement, with suggestions that this may have been avoided if they had been better supported following the death.

> *‘Work was incredibly unsupportive…. I was judged in a negative light as my performance dropped immediately after the funeral. They were also annoyed I took some time off. They had only offered one day of compassionate leave for the funeral. I feel v. strongly that companies need to [review] their bereavement policies.’ (Bereaved daughter, BRID470)*

Some people were disinclined to take time off at a time of financial uncertainty, from fear of losing their jobs. Those running their own businesses described additional financial pressures and concerns over loss of livelihood. Where people had experienced job-loss, the negative impact on their mental health was noted. People in frontline jobs described difficulties managing their grief and working in pressured, public-facing roles. Others described the isolating effects of being furloughed or working remotely, which made it harder for them to connect with and feel supported by their colleagues. Workload pressure also prevented people taking time off or seeking the support that they needed. Healthcare workers explained that their clinical work meant that they were constantly reminded of the ongoing threat of the pandemic and the personal trauma they had suffered, but that they also felt guilty about taking time off and being unable to support their colleagues.

> *‘We have had to seek out legal support as my husband was unfairly dismissed from work during the pandemic. This has caused us a lot of stress. It would have been very useful to have access to someone who was able to tell us what we were legally able to pursue early on, as my husband’s mental health spiralled out of control due to these additional stressors.’ (Bereaved mother, BRID267)*

#### Support from the health and social care system

Difficulties accessing bereavement support from GPs and bereavement services were commonly reported (and described in detail elsewhere, Harrop et al 2021). Difficulties relating to support from other parts of the health and social care system were also identified, including help with the effects of long COVID and managing other chronic conditions alongside their grief. A participant who experienced pregnancy loss described the trauma of her experience. She stated that her anxiety was exacerbated by being without her partner during scans, consultations and surgery, as well as a lack of support afterwards due to the focus on Covid-19. Many participants felt let down by absent or inadequate follow-up contact after the death from GPs, hospitals and care homes. They described feeling lost in having to manage their bereavement alone, as well as not knowing where to turn to receive answers or further support. The need for timely provision of verbal and written information relating to bereavement services, registering the death and arranging funerals was also identified.

> *‘What would have helped: a) asked if I was okay b) was there anyone that I was close with that I could talk to (family, friends, partner etc.) c) put me in touch with some bereavement support, either in person, online or via the phone (preferably send these to my email so that I could look at them later - my mind was so scrambled I couldn’t barely remember anything) d) clarify what happens next e.g. death certificate, registering death, organizing funeral etc.’ (Bereaved granddaughter, BRID486)*

### The Dual Process Model: Exacerbation of Loss and Restoration Oriented Coping

Following the inductive analysis presented above, these themes were considered using the DPM as an analytic lens and mapped to the lists of pandemic-specific loss- and restoration-oriented stressors and reactions developed by Stroebe and Shut (2021). Full results are provided in Tables 3 and 4 (supplementary file 1), confirming and expanding upon many of the factors in these lists. Examples of loss-oriented (LO) stressors included limited contact at the end of life and restricted funeral and memorialisation practices, leading to LO reactions of guilt, anger and problems accepting the death. Examples of restoration-oriented (RO) stressors and reactions included severely curtailed social networks, support systems and social/recreational activities, all of which impacted peoples’ ability to cope. New additions from our analysis include the acute stress and distress caused by difficulties with death administration (RO), more extensive sets of workplace pressures and strains (RO), lack of time and space to grieve (LO) and find respite (RO), and reluctance to seek support due to perceived widespread suffering, empathy fatigue and lack of understanding from those not sharing these ‘exceptional’ experiences (RO).

## DISCUSSION

The findings of these two national UK surveys demonstrate the profound and wide-reaching impact that the pandemic has had on the lives of people who have been bereaved, providing qualitative evidence for the many challenges of pandemic bereavement. These include traumatic death experiences, restricted memorialisation practices and contending with the ongoing threat of the virus, as well as societal responses to the pandemic. Severely curtailed social networks, difficulties at work and with accessing help from health and social care professionals have further undermined people’s ability to cope with and adapt to bereavement at this time. Almost all of our themes could be mapped to the lists of pandemic LO and RO stressors/reactions developed by Stroebe and Schut (2021), with several new additions also identified. These results demonstrate the utility of using the DPM as an analytic lens for conceptualising pandemic bereavement experiences (Stroebe and Shut 1999), as well as the important evidence-based contributions of this study to this framework.

We found extensive examples of LO stressors. Consistent with other research findings, these included, lack of contact with dying relatives/friends and difficulty saying goodbye due to visiting and lockdown restrictions, communication difficulties with healthcare providers at the end of life (Becque et al. 2021, Hanna et al. 2021, Mayland et al. 2021) and the sudden and unexpected nature of many deaths. The high prevalence of these experiences were confirmed in the quantitative results of the same study, with greater occurrence of several of these problems observed for Covid-19, hospital and care-home deaths (Selman et al. 2021c). Other LO stressors, not evidenced in previous research, include the profound changes in funeral practices and the prohibition of traditional gatherings held around the time of the funeral. The lack of opportunity for physical contact with family and friends and severely disrupted support networks also meant that people were unable to grieve together and to remember their relative/friend collectively.

Loss oriented reactions included people feeling guilty that they let their relatives down at the end of life, as observed in another UK study (Hanna et al 2021, Mayland et al 2021), as well as in their funeral and mourning arrangements. Anger was also felt towards hospital policies and practices and more generally at the public and governmental response to the pandemic; reflecting similar themes to those identified in a recent analysis of social media commentary amongst bereaved relatives (Selman et al. 2021a). Relatives were frequently left with unanswered questions and with niggling doubts, which made it harder for them to process and reconcile their feelings surrounding the death. The significance of this reaction is reflected in the quantitative survey data: 60% of participants experienced high or fairly high needs for help ‘dealing with my feelings about the way my loved one died’ (Harrop et al 2021). Grieving in isolation, without recourse to usual rituals and practices, was also felt to make the deaths seem less real and made it harder for people to find closure and begin to grieve. These reactions are consistent with the quantitative findings of another pandemic study; that ‘disrupted meaning’ contributed to worse grief outcomes (Menzies et al. 2020; Breen et al 2021). Other LO reactions, not previously documented, related to the intensity of family life and home-schooling during lockdown, which denied parents in particular the time and space needed to grieve and process their loss. We also observed the distressing, dehumanising effects of the mass bereavement context and associated media coverage, the negative content of which is documented in analyses of media coverage of bereavement during the pandemic (Selman et al 2021b; Sowden et al. 2021). People not only faced constant reminders of their loss and trauma but also perceived a societal devaluing of life and death.

Examples of RO stressors and reactions not previously evidenced in pandemic studies included high levels of stress and distress caused by difficulties dealing with death-related administration and sorting out the affairs of the deceased person. Although not as prevalent as the emotional support needs identified in this study, around a quarter of participants reported high/fairly high needs for help with administrative tasks and accessing financial and legal information and advice (Harrop et al. 2021). The alienating effects of social division and disharmony caused by public and officials’ responses to the pandemic, including the sharing of conspiracy theories and disregard of safety regulations, presented further challenges to restorative processes. People also felt less comfortable seeking help due to concerns over burdening others at this universally stressful time, perceived empathy fatigue and a sense that others could not understand the unique grief that they were experiencing.

Severely diminished support networks during lockdown periods, as well as fear and self-imposed isolation in response to the ongoing threat of the virus (particularly amongst those bereaved by Covid-19) meant that opportunities to engage in social activities were limited, as were other usual coping or recreational activities. This not only meant that people were denied the emotional support that they needed to help them cope (Harrop et al 2021), but also that they were prevented from finding new meaning, purpose or respite from their grief; difficulties compounded by added family pressures and responsibilities during lockdowns. These challenges were confirmed in the quantitative analyses, which found that around a half of people experienced high/fairly high needs for help with ‘loneliness and isolation’, ‘feeling comforted and reassured’, ‘finding balance between grieving and other areas of life’ and ‘regaining sense of purpose and meaning in life’ (Harrop et al. 2021). Social isolation and loneliness were especially prevalent among bereaved partners, as might be expected, but also people bereaved by COVID-19 deaths (Selman et al. 2021c), likely reflecting the social consequences of Covid bereavement described here. Difficulties relating to workplaces and employers, such as lack of bereavement leave, compassion or understanding, enhanced isolation due to furlough and homeworking and job and financial insecurities, made this process of adaptation all the more challenging. Recovery was further limited by difficulties accessing bereavement support services and getting help from other parts of the health and social care system (Harrop et al 2021; Selman et al 2021c).

### Strengths, limitations and implications for research

Both surveys benefit from the longitudinal nature of their design: the BeCOVID project is the first to longitudinally investigate peoples’ experiences of bereavement support during the Covid-19 pandemic in the UK and the COPE study is one of the largest in-depth studies of public experiences and perceptions during the UK pandemic. The COPE survey was restricted in the number of questions relating to bereavement due to the module being only one of many within the overall survey. We were therefore unable to collect as much detailed information related to when and where the deaths occurred, experiences at the end of life and perceptions of bereavement support among COPE participants, and the data were not as comprehensive or detailed as in the BeCOVID study.

Although both study sample sizes are large the COPE study recruited most participants through HWW, under-representing other UK nations. In addition, people from minority ethnic backgrounds and men are underrepresented in both data sets. Recruitment was predominantly online which meant we were less likely to reach the very old or other digitally marginalised groups. Understanding the experiences of these groups, who are likely to have experienced greater vulnerability and challenges during their bereavement, is an important area for future research. This paper has focused on the challenges of pandemic bereavement. Forthcoming publications will focus on what has helped bereaved people cope and adapt during the pandemic with messages for improving the resilience and support available to them.

## Conclusion

These findings demonstrate the exceptionally difficult sets of experiences associated with bereavement during the pandemic, defined by significant disruption to end-of-life, death and mourning practices, as well as social support networks and services. We have shown the DPM provides a useful framework for conceptualising the additional challenges associated with pandemic bereavement and their impacts on grieving, coping and mental health.

Based on these findings we make six recommendations for improving the experiences of people bereaved during and following this and future pandemics:

1. Taking steps to reduce the trauma associated with death experiences, through improved communication with and involvement of families (Selman et al. 2021c), safe facilitation of family visiting to healthcare settings, and, where this is not possible, connecting families and loved ones through accessible remote communication methods.
2. Healthcare providers improving family support after a death, including routinely providing opportunities to discuss patient care and the circumstances of the death, and information about locally and nationally available bereavement support.
3. Strengthening the bereavement support sector, including greater resourcing and expansion of national support, regional services in areas with long waiting lists and strategies to improve awareness of bereavement support options (Harrop 2021).
4. Tackling loneliness and social isolation, including flexible support bubble arrangements for the recently bereaved when restrictions are in place and informal community-based interventions aimed at strengthening social networks, grief literacy and communication skills, as championed by compassionate communities networks (Breen et al. 2021; Compassionate Communities UK; Harrop et al. 2021).
5. Developing, promoting and adhering to guidance and best-practice recommendations regarding: a) funeral options (including virtual) during times of social restrictions, b) supporting those administering the death of their deceased relative, and c) supporting bereaved employees (e.g. see <u>Managing bereavement in the workplace – a good practice guide (acas.org.uk)</u>.
6. Providing opportunities for remembrance, greater respect and listening to those bereaved. This requires media recognition of the dehumanising impacts of death-statistics, the need to give voice to the stories of the bereaved and provide more supportive narratives (Snowden et al. 2021; Selman et al 2021b); national and local initiatives which support private and public remembrance, such as dedicated spaces/ memorials and national days of reflection; and inclusive consultation with those recently bereaved (e.g. see <u>UK Commission on Bereavement (bereavementcommission.org.uk)</u> to ensure lessons are learned for future pandemics.

## Data Availability

Full data sets will be made available following study closure in February 2022. Data sharing requests will be considered prior to this and should be directed to Dr Emily Harrop

## Acknowledgements

Our thanks to everyone who completed both surveys for sharing their experiences and voluntarily providing their time for these studies, and to all the individuals and organisations that helped disseminate the surveys. We are grateful to our patient and public involvement members for their invaluable contributions to designing and steering this research. We would also like to thanks all collaborators and advisory group members for both studies.

The COPE study was facilitated by HealthWise Wales, the Health and Care Research Wales (HCRW) initiative which is led by Cardiff University in collaboration with the SAIL Databank, Swansea University. We are grateful for their invaluable support and expertise. We would like to thank Cardiff Metropolitan University, Cardiff University, PRIME Centre Wales, and Swansea University who have all been immensely supportive of this work, allowing out team the time and providing infrastructure to get the study up and running quickly during the very early stages of the pandemic.

## Author contributions

E.H. and L.E.S. designed the BeCovid study, led the application for funding and are co-principal investigators; A.T.B. and E.H. drafted the paper; M.L., A.B., D.F., E.C. E.S., A.N., A.T.B., K.S., S.G. are members of the BeCovid research team or study advisory group and contributed to the design of the study and survey. R.P. is principal investigator on the COPE Study. R.P and E.H designed the bereavement module in this survey. E.H., A.T.B., K.B., E.S. and S.G. conducted the thematic analysis of qualitative data. All authors contributed to drafting the paper and read and approved the final manuscript.

## Ethical approval

The BeCovid study protocol and supporting documentation was approved by Cardiff University School of Medicine Research Ethics Committee (SMREC 20/59). The study was conducted in accordance with the Declaration of Helsinki and all respondents provided informed consent at the beginning of the baseline survey and provided consent to re-contact at the end of the survey.

Ethical approval was obtained for the COPE Study from the Cardiff Metropolitan University Applied Psychology ethics panel on 13.3.20 (Project reference Sta-2707). Participants provided consent and confirmed eligibility electronically at the beginning of the baseline survey and provided consent to re-contact at the end of the survey.

