## Supplementary File 1 for "“It was brutal. It still is”: A qualitative analysis of the challenges of bereavement during the COVID-19 pandemic reported in two national surveys"

**Supplementary File One**

Table 3 Comparison of stressors in this analysis with the DPM Pandemic Framework (Stroebe and Schut 2021)

| LO Stressors from Strobe & Shut framework | Related LO Stressors from BeCovid & COPE data | RO Stressors from Strobe & Shut framework | Related RO Stressors from BeCovid & COPE data |
|---|---|---|---|
| Lack of emotional & practical preparation time | -Sudden and unexpected nature of Covid-19 deaths | – Loss of work, unemployment, livelihood/financial insecurity, furloughs, salary reductions, homelessness | Job loss/ financial worries |
| Circumstances of terminal illness, dying process (e.g., isolation, desolate dying situation; patient's physical discomfort, difficulty breathing; illness-related uncertainty) | -Perceived patient suffering | Lockdown-related difficulties with one's work, harder to get new employment if needed post-loss | • Isolation caused by working from home, enforced absence/separation from colleagues due to furlough.<br>• **Pressure to return before ready, exacerbated by fear of job loss or acute workloads of frontline staff*** |
| The traumatic burden for (bereaved) relatives of treatment of their dying relative in isolated ICU conditions | Died alone<br>Perceived lack of dignity | Medical support concerns (e.g., lack of insurance; health care access; failed medical intervention due to lack of resources, facilities, means to pay) | Limited availability of/access to bereavement services |
| Difficulty/impossibility to say goodbye due to enforced isolation; realization of death more difficult without viewing the body or saying goodbye, associated with denial and avoidance of the loss) | -Myself or other family unable to visit/say goodbye<br>- No change of clothes/preparing the body/seeing body<br>- Difficulty processing/accepting death | Care, management and treatment for existing mental and physical illnesses (associated with accentuation of health care disparities) | Difficulties managing other health conditions/accessing services |
| Morally distressing circumstances: perceptions of violation to one's own moral or ethical code (relating to suboptimal care of the dying person, due to resource scarcity (including difficulties relating to ethical decisions in limited resources) | Perceived poor treatment/care decisions at End of Life | High-risk frontline, repeated exposure stress; own COVID-19 illness, intensive care admission | Pressures on frontline staff- feeling that can't take time off, constant reminders of own trauma (clinical staff) |
| Circumstances of death unexpected and/or traumatic (associated with continued confrontation, rumination and complicated grieving, but note avoidance too). | -Sudden/unexpected deaths<br>-Unable to say goodbye or visit | Quarantine/confinement/social isolation (restrictions in social, recreational, and occupational activities, including travel e.g., for attending memorial ceremonies | Forced isolation (covid)<br>Unable to attend memorial ceremonies (myself or close family) |

| | | | |
|---|---|---|---|
| Experience of "staggering", multiple losses/deaths of close persons and their ongoing nature; threat to oneself (particularly. among vulnerable older, sick) | -Other recent/multiple bereavements | Family (particularly children) and community level difficulties/disruptions in living arrangements (e.g., schooling irregularities: home schooling; access to shops) | • Family responsibilities and pressures during lockdown e.g. home-schooling<br>• **Lack of time and space to take a break and find respite from grief*** |
| Frequent reminders of death (rates), media coverage, exposure to distressing images; hearsay, potentially impacting negatively on adaptation | Constant reminders in media coverage | Painful physical separation from family and close friend | Grieving difficult without physical contact and comfort |
| Pandemic circumstances may be dealt with by suppression of emotions (in certain cultures particularly), linked to avoidance and denial rather than confrontation | | Family tensions and quarrels (violence) associated with staying at home, no family/friends visits | Strained relationships |
| Profound changes in funeral practices, severe restrictions (inc. social & relational ones): enforced direct cremations; banned ceremonial attendances | Restriction to numbers<br>Service features (singing, flower, no service, locked cemetery) | Disruption of connectedness, autonomy and freedom through suspension of gatherings, travel, closing of religious centres, lockdowns, social distancing | Feelings of loneliness and isolation |
| Lack of opportunity for physically given/received support | Grieving in isolation | Erosion of coping resources (social support), relating to social isolation, (lack of positive appraisal: sense of community, uplift). | • Lack of social support, due to distancing and **perceived lack of empathy or understanding relating to exceptional nature of pandemic bereavement and widespread societal burden***<br>• Disrupted coping mechanisms- opportunities for respite and distraction |
| Lack of social/cultural recognition of the loss, related to impaired support resources; absence of the soothing rituals with opportunity to express grief/comfort | Isolated from support networks at end of life, death and soon after<br>Disrupted mourning practices | Needed support for vulnerable bereaved people may be lacking due to distancing and withdrawal | • Grieving and coping difficult without physical contact and comfort from close others<br>• Feelings of loneliness and isolation |
| | **Lack of time and space to grieve (e.g due to homeschooling) *** | (Perceived) rejection (e.g., stigmatization from others due having a COVID-19 death in the family) | |
| | | Impact of negative campaigns, victimization, rise in racism | • Conspiracy theories<br>• Concern over disproportionate impact on BAME communities/health inequalities |

| | | | |
|---|---|---|---|
| | | Disruptions to social norms, face-to-face rituals, mourning practices impacting on ongoing life beyond grief, grieving | • Isolated from support networks at end of life and early bereavement<br>• Grieving difficult without physical contact and comfort |
| | | Worries, fears about getting the viral infection, fear of contamination/contaminating others | Fear for myself and others catching or dying of covid |
| | | Difficulties dealing with new technology needed due to distancing (online medical forms, phone appointments, etc.) | Difficulties with death related administration during lockdown |
| | | Uncertainty about the future (under pandemic circumstances) | |
| | | Lack of/disruption of routines, dislocation, loss of pre-crisis ways of life, threatened loss of hopes and dreams for the future, shattered assumptions about normal life and connections with the world | Disrupted coping mechanisms e.g difficulty engaging in previous or new activities, getting out the house |

***new additions to the DPM pandemic framework identified from BeCovid and COPE data analysis**

Table 4 Comparison of reactions in this analysis with the DPM Pandemic Framework (Stroebe and Schut 2021)

| **LO Reactions from Strobe & Shut framework** | **Related LO Reactions from BeCovid & COPE data** | **RO Reactions from Strobe & Shut framework** | **Related RO reactions from BeCovid & COPE data** |
|---|---|---|---|
| | Feelings of guilt/that let loved one down | A rise in level of general anxiety, worries | • **Societal strains of pandemic; e.g. Not wanting to burden others at time of universal suffering, perceived lack of empathy***<br>• Feeling generally overwhelmed |
| Anger toward system/officials about the death: "undignified" body disposal; regarding the patient's own mistakes; | Anger at government handling of pandemic<br>Anger relating to hospital policies and practices at EoL (e.g. patient care, family visiting) | Feelings of lack of safety, insecurity | • Fear for myself and others catching or dying of Covid. |
| Moral responsibility: guilt, anger, anguish, distress exacerbated by feelings of moral responsibility | Feelings of guilt/ that let loved one down | Experiencing prejudice against oneself | |
| Loneliness; excessive bitterness and a sense of perceived purposelessness in life. | Feelings of loneliness and isolation/ diminished meaning and purpose relating to loss of relationship with deceased | Feeling upset related to an intolerance of uncertainty | |
| Anticipatory grief phenomena, which are also threatening, given uncertainty of illness outcome | | Loss of feelings of control or purpose | Diminished meaning and sense of purpose related to lack of social/recreational etc activities to occupy oneself with<br>Disrupted coping mechanisms<br>Finding it hard to make decisions |
| – Disenfranchised grief: 1. Too many COVID-19 deaths for individual recognition, the deceased considered a statistic, causing complications in grieving 2. Linked to failure by deceased to follow mandated rules, impacting on social interactions in bereavement | Dehumanising effect of mass bereavement death as a statistic | Feeling trapped through isolation | Feelings of loneliness and isolation/unable to move forwards |
| | | Feeling vulnerability to the spread of rumours, misinformation | Alienation and distress related to social discord/conspiracy theories/**others disregard of Covid regulations*** |
| | | Fears for one's own mortality | Fear for myself and others catching or dying of covid |

| | | | |
|---|---|---|---|
| | | Experiencing the allocation of blame toward oneself, a victim of discrimination or of hate crimes | |
| | | | **Acute stress/distress experienced relating to difficulties dealing with death administration*** |

***new additions to the DPM pandemic framework identified from BeCovid and COPE data analysis**
